# Striatal dopamine gene network moderates the effect of early adversity on the risk for adult psychiatric and cardiometabolic comorbidity

**DOI:** 10.1101/2022.04.23.22274209

**Authors:** Barbara Barth, Danusa Mar Arcego, Euclides José de Mendonça Filho, Randriely Merscher Sobreira de Lima, Carine Parent, Carla Dalmaz, André Krumel Portella, Irina Pokhvisneva, Michael J Meaney, Patricia Pelufo Silveira

## Abstract

Cardiometabolic and psychiatric disorders often co-exist and share common early life risk factors, such as low birth weight. However, the biological pathways linking early adversity to adult cardiometabolic/psychiatric comorbidity remain unknown. Dopamine (DA) neurotransmission in the striatum is sensitive to early adversity and influences the development of both cardiometabolic and psychiatric diseases. Here we show that a co-expression based polygenic score (ePGS) reflecting individual variations in the expression of the striatal dopamine transporter gene (*SLC6A3*) network significantly interacts with birth weight to predict psychiatric and cardiometabolic comorbidities in both adults (UK Biobank, N= 225,972) and adolescents (ALSPAC, N= 1188). Decreased birth weight is associated with an increased risk for psychiatric and cardiometabolic comorbidities, but the effect is dependent on a striatal *SLC6A3* ePGS, that reflects individual variation in gene expression of genes coexpressed with the SLC6A3 gene in the striatum. Neuroanatomical analyses revealed that SNPs from the striatum *SLC6A3* ePGS were significantly associated with prefrontal cortex gray matter density, suggesting a neuroanatomical basis for the link between early adversity and psychiatric and cardiometabolic comorbidity. Our study reveals that psychiatric and cardiometabolic diseases share common developmental pathways and underlying neurobiological mechanisms that includes dopamine signaling in the prefrontal cortex.

## Introduction

The co-occurrence of more than one chronic disease^1^ has high prevalence in primary care settings^2^, inflating health care utilization and functional disability^3^. Psychiatric and cardiometabolic disorders, which are highly comorbid^4,5^, rank amongst the leading global causes of disability-adjusted life years worldwide^6,7^. Prospective studies show a bi-directional relationship between psychiatric and cardiometabolic conditions^8^. Meta-analytic evidence from longitudinal studies indicates that diabetes increases the risk for depression by approximately 25% and that depression increases the risk for type 2 diabetes by 40-60%^7,9^. The odds for depression also increase with one or more non-psychiatric coexisting chronic conditions, especially coronary artery disease, chronic arthritis, and stroke^10^. Anxiety is also associated with 41% increased risk of developing cardiovascular disease^11^. Among adult patients with schizophrenia, the prevalence of diabetes averages of 15%, which is higher than the 10% prevalence of diabetes in the general population. This association persists even after controlling for obesity and use of antipsychotic drugs^12^.

The underlying mechanism for these comorbidities remains unknown, but an emerging explanation is that psychiatric and cardiometabolic disorders share common developmental pathways. For example, low birth weight broadly reflects an unhealthy fetal environment and is considered a prevalent form of early life adversity^13^ associated with increased morbimortality throughout the life course^14^ and is specifically linked to cardiometabolic^14–18^ and psychiatric disorders^19–24^. An obvious question concerns the biological mechanisms that underlie such a developmental trajectory involved in the development of cardiometabolic and psychiatric comorbidities.

The brain dopamine system is highly sensitive to early adversity^25^ and proposed as a mechanism underlying developmental pathways to multiple psychiatric and metabolic comorbidities^26,27^. Early life adversity such as fetal growth restriction that leads to low birth weight alters dopaminergic signaling ^28–30^. Dysfunction of dopamine neurotransmission in both the ventral and dorsal striatum associates with depression^31^, as well as dysregulated food intake and altered energy homeostasis^29,30,32^. Striatal dopamine signaling also appears to regulate systemic glucose metabolism in humans.^33^ The striatum harbors dopaminergic neurons^34^ and the striatal dopamine transporter (DAT) is a critical regulator of striatal dopamine release and reuptake.^35^ Dopamine signaling is influenced by core metabolic hormones such as leptin and insulin, through their actions on the expression and function of DAT ^32,36^, which is encoded by the *SLC6A3* (solute carrier family 6 member 3) gene.

Based on the large evidence supporting the relation between metabolism, mental health and striatal dopaminergic neurotransmission, as well as the effects of early adversity on striatal dopamine function, we hypothesized that the striatal *SLC6A3* gene network underlies the association between early life adversity and the comorbidity between psychiatric and cardiometabolic disorders in humans. We therefore aimed to test if individual differences in the function of a striatal *SLC6A3* gene network might moderate the effects of early life adversity on psychiatric and cardiometabolic comorbidities in adults and adolescents. To achieve this, we created a *SLC6A3* striatal co-expression-based polygenic score (striatum *SLC6A3* ePGS) reflecting the genetic capacity for expression of the striatal DAT1 gene network (possibly influencing dopamine signaling) and analyzed the effect of its interaction with birth weight on the comorbidity of psychiatric and cardiometabolic conditions in adults (UK Biobank) and adolescents (Avon Longitudinal Study of Parents and Children, ALSPAC).

## Methods

### Participants

We used genomic and phenotypic data from two cohorts, one from adults (Uk Biobank), and one from adolescents (Avon Longitudinal Study on Parents and Children, ALSPAC). **Adult cohort**: The UK Biobank is a large population-based study from the United Kingdom^37^. Participants, aged 37-73, were recruited between 2006 and 2010 resulting in 502,543 subjects. Detailed description of the inclusion/exclusion criteria for the current analysis and the corresponding sample size at each step can be found in *Supplementary information***, Supplementary** Figure 1. After all exclusions and inclusion criteria, the number of subjects that remained for the analysis was 225,972 (mean age = 55.22, SD = 8.08) (**Table 1**). We used all the data available for the brain imaging analysis considering the inclusion/exclusion criteria (*Supplementary information***, Supplementary Figure 1**, N=11,167, mean age = 53.86, SD = 7.39).

**Table 1.** Description of the baseline characteristics in UK Biobank sample for high and low striatum *SLC6A3* ePGS groups, which were defined by median split of the ePGS score.

| Characteristics | Low ePGS |  | High ePGS |  | P value |
| --- | --- | --- | --- | --- | --- |
|  | Mean or % | N or SD | Mean or % | N or SD |  |
| Sex - Male | 39.2% | 44324 | 39.5% | 44615 | 0.21 |
| Birth weight (grams) | 3314 | 658 | 3321 | 659 | <0.001 |
| Breastfed as a baby | 70% | 70838 | 69.5% | 70310 | 0.03 |
| Maternal smoking around birth | 28.9% | 29209 | 29% | 29222 | 0.71 |
| Completed full-time education at 14-years of age or younger | 1.2% | 837 | 1.2% | 830 | 0.82 |
| Age at recruitment (years) | 55.17 | 8.09 | 55.26 | 8.07 | 0.01 |
| Townsend deprivation index at recruitment | -1.44 | 3.01 | -1.52 | 2.95 | <0.001 |
| Current or previous smoking | 43.4% | 48829 | 43.3% | 48738 | 0.696 |
| BMI at recruitment | 27.25 | 4.88 | 27.28 | 4.86 | 0.264 |
| ICD10 F10-F19 Mental and behavioural disorders due to psychoactive substance use | 4.2% | 4694 | 4.1% | 4584 | 0.25 |
| ICD10 F20-F29 Schizophrenia, schizotypal and delusional disorders | 0.2% | 266 | 0.3% | 290 | 0.33 |
| ICD10 F30-F39 Mood [affective] disorders | 0.2% | 266 | 0.3% | 290 | 0.33 |
| ICD10 F40-F48 Neurotic, stress-related and somatoform disorders | 2.5% | 2772 | 2.5% | 2769 | 0.98 |
| ICD10 E11 Non-insulin-dependent diabetes | 4.7% | 5257 | 4.4% | 5012 | 0.01 |
| ICD10 I70 Atherosclerosis | 0.3% | 316 | 0.3% | 316 | 1 |
| ICD10 I63 Cerebral infarction | 0.8% | 879 | 0.8% | 879 | 1 |
| ICD10 I20-I25 Ischaemic heart diseases | 6.9% | 7759 | 6.8% | 7630 | 0.29 |
| Birth weight (<= 2.5kg, > 2.5kg) | 89.7% | 101344 | 90% | 101665 | 0.03 |

Townsend deprivation index in UK Biobank: A measure of the level of social deprivation that a person lives in. The index was calculated based on previous national census. Participant is given a score reflecting the output area in which their postal code is located. Four key aspects are considered in the index: the percentage of unemployment, overcrowded households, households without a car and non-home ownership.

### Adolescent cohort

To explore our findings in an earlier developmental time point we used data from the Avon Longitudinal Study of Parents and Children (ALSPAC) cohort^38–40^. This is a transgenerational prospective observational cohort that recruited 14, 541 pregnant women residents in Avon County, UK. Additional recruitment (N=913) was performed later during Phases II, III and IV respectively, bringing the total sample size of prospective mother-child dyads to 15,658. For more information on ALSPAC variables, please see http://www.bristol.ac.uk/alspac/researchers/our-data/. Data from the adolescent offspring aged between 15.5 and 17.5 were used in this study. Only subjects with available phenotypic data of interest, early life adversity measure, in this case birth weight and genotyping data were considered for the analyses (N=1,188) (**Table 2**).

**Table 2.**
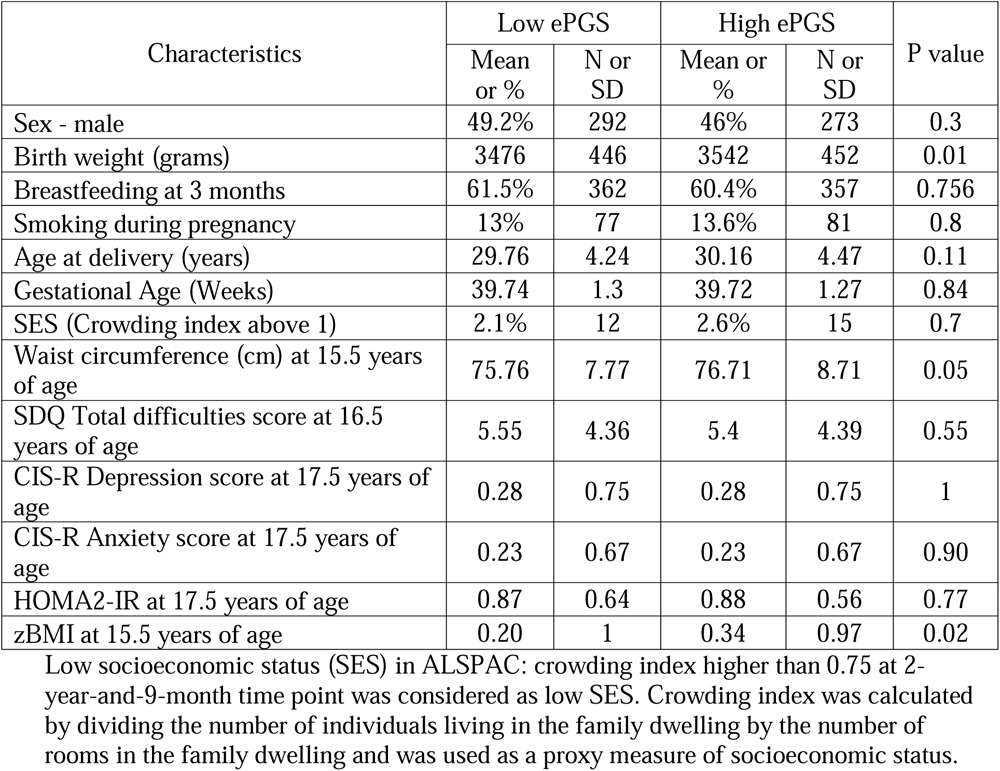
Description of the baseline characteristics of the ALSPAC sample used according to high and low striatum *SLC6A3* ePGS. Groups were divided by the median of the ePGS score.

Detailed description of the inclusion/exclusion criteria and the corresponding sample size at each step can be found in *Supplementary information***, Supplementary Figure 2**. See *Supplementary Information* Supplementary Methods for detailed description of the genotyping procedure for each cohort.

### Ethics approval and consent

#### UK Biobank

Informed consent was obtained from each participant, and the project has been approved by the North-West Multicentre Research 580 Ethics Committee (REC reference 11/NW/0382), the National Information Governance Board for Health and Social Care, and the Community Health Index Advisory Group for UK Biobank.

Consenting participants provided baseline information, answered questions, had measurements and biological samples collected. This research has been conducted using the UK Biobank Resource under application number 41975.

#### ALSPAC

Participants provided informed written consent to participate in the study. Ethics approval for the study was obtained from the ALSPAC Ethics and Law Committee and the local research ethics committees (a full list of the ethics committees that approved different aspects of the ALSPAC studies is available at http://www.bristol.ac.uk/alspac/researchers/research-ethics/). Consent for biological samples has been collected in accordance with the Human Tissue Act (2004).

Consent for publication was obtained from UK Biobank and ALSPAC management teams. The use of these datasets was locally approved by the Centre intégré universitaire de santé et de services sociaux de l’Ouest-de-l’Île-de-Montréal Research Ethics Board under application number IUSMD-21-73.

### Identification of the striatal *SLC6A3* co-expression gene network and ePGS calculation

Figure 1A shows the steps involved in the identification of the gene co-expression networks and the calculation of the ePGS score. The ePGS was calculated considering genes co-expressed with the *SLC6A3* gene in the striatum. As described previously^41–48^, we began by using brain region-specific RNA sequencing data from mice available at GeneNetwork (http://genenetwork.org/, HBP Rosen Striatum M430V2 (Apr05) RMA Clean)^49^ to identify *SLC6A3* co-expressed genes (absolute value of co-expression correlation with *SLC6A3* gene greater or equal to r= 0.5). GeneNetwork was used to obtain gene expression from rodents since our previous findings demonstrated multiple effects of early life adversities, especially poor fetal growth, on dopaminergic mesocorticolimbic system in rodents^28–30,50–54^. We then converted *SLC6A3* co-expressed genes to human orthologs by using the biomaRt package^55^. Since we were interested in gene networks that were active during the early developmental period in which adversity occurred and when the brain is still undergoing core maturational processes in humans, we used BrainSpan to select autosomal transcripts expressed at least 1.5-fold more during fetal and childhood periods (0–60 months after birth) in comparison to adulthood (20–40 years of age). This process resulted in a list of striatal *SLC6A3* co-expressed genes. We then mapped all the existent SNPs in the human ortholog genes comprising the striatum *SLC6A3* gene network using biomaRt package^55^ in R and gathered all gene-SNP pairs from the GTEx dataset in human striatum. These lists were merged with the genotyping data from UK Biobank and ALSPAC cohorts, respectively, retaining only common SNPs and subjecting the final SNP lists to linkage disequilibrium clumping (r2 < 0.2) within 500kb radious to eliminate redundant SNPs. The process resulted in 1532 independent functional SNPs retained in UK Biobank and 1663 SNPs in ALSPAC. The final score included 67 genes in our discovery sample (UKB) (*Supplementary information***, Supplementary Table 1**).

**Figure 1.**
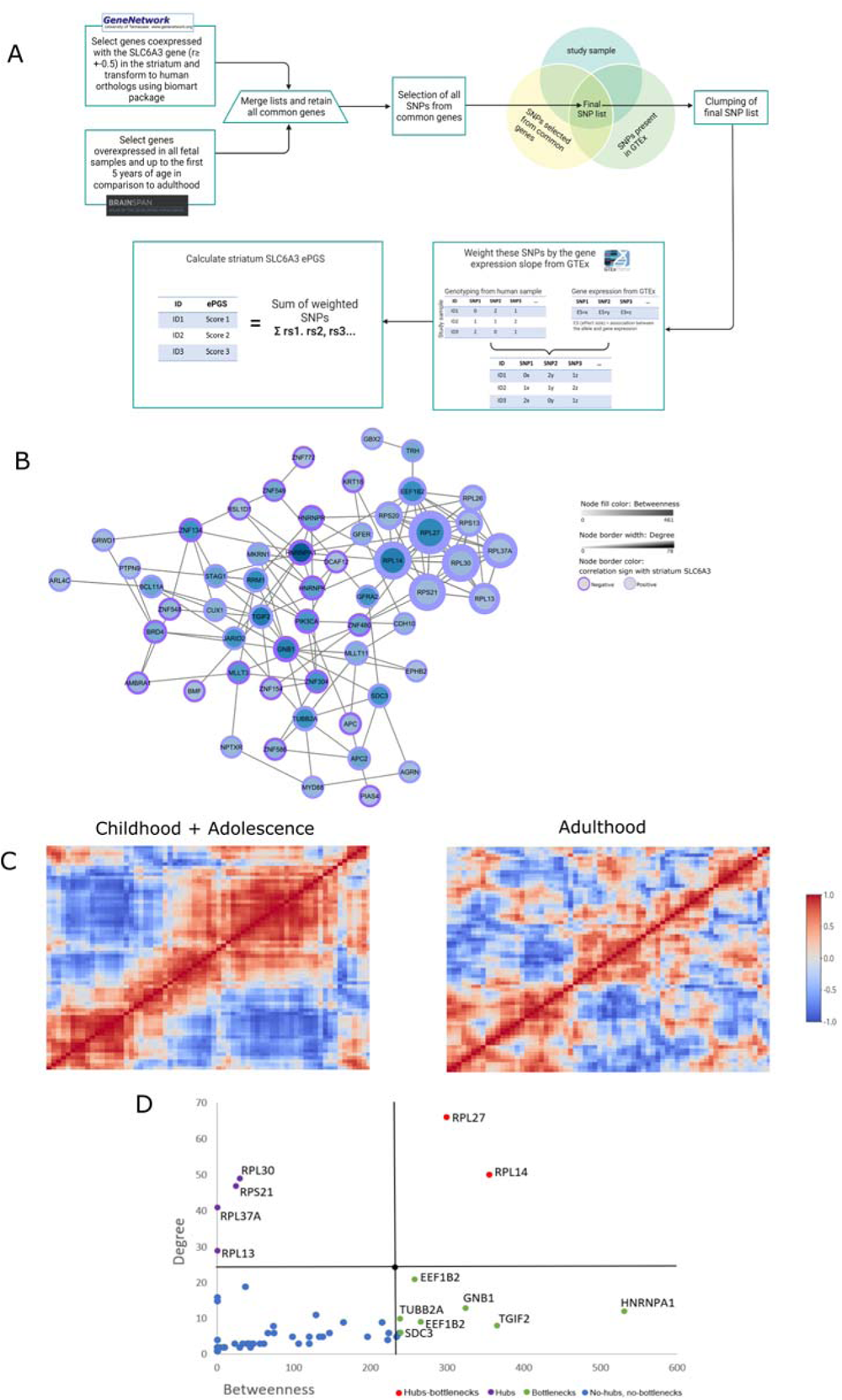
Construction and characterization of the striatum *SLC6A3* gene network. A, calculation of the expression based polygenic score (ePGS) from the genes co-expressed with the *SLC6A3* gene in striatum. GeneNetwork was used to generate a list of genes co-expressed with *SLC6A3* in striatum in mice, which were then converted to human orthologs. BrainSpan was used to identify genes overexpressed within striatum in fetal samples and up to 5 years of age in comparison to adult samples. All SNPs from these genes, common between the study sample and GTEx databases, were retained and included in the final list of SNPs. This final list was subjected to linkage disequilibrium clumping, with removal of highly correlated SNPs. Next, for each SNP, a number of alleles at a given SNP (rs1, rs2…) was multiplied by the estimated effect of the genotype-gene expression association from GTEx. The sum of these values over all SNPs provides the striatum *SLC6A3* ePGS. B, striatum *SLC6A3* ePGS co-expression gene network. Co-expression pattern was mined from GeneMANIA^61^. The color of the node border represents the correlation sign with the *SLC6A3* gene according to GeneNetwork co-expression matrix (dark purple represents negative and light purple positive correlation). Node color intensity represents betweenness (number of times a node acts as a bridge between nodes). Node border width represents the number of connections a node has with other nodes (total degree). C, co-expression of genes included in the striatum *SLC6A3* gene network in humans at different ages according to BrainSpan. D, topological properties of the striatum *SLC6A3* gene network, showing hubs (with degrees higher than +1SD above mean), bottlenecks (betweenness higher than +1SD above the mean), and hub-bottlenecks. Lines in black indicate mean + 1 SD for degrees and betweenness. Hub and hub-bottleneck genes are related to ribosomal structure. Among the bottleneck genes, HNRNPA1 is involved in the packaging of pre-mRNA into particles and transport from the nucleus to the cytoplasm, as well as splicing. The SDC3 gene may play a role in cell shape organization and has been associated with obesity^74^.

To calculate the striatal *SLC6A3* ePGS, the number of effect alleles (genotype information from the study samples) at a given SNP was weighted using the estimated brain-region-specific effect of the SNP on gene expression from the GTEx data^56^. We also accounted for the direction of the co-expression of each gene with *SLC6A3* in the network, by multiplying the weight by -1 in case the expression of a gene was negatively correlated with the expression of the *SLC6A3* gene in the network – therefore, the higher the score, the higher the expression of the genes that compose the network. The sum of the weighted values from all SNPs for each individual in the cohorts resulted in the region-specific striatal scores. The striatal *SLC6A3* expression-based polygenic score (ePGS) is a continuous measure that reflects variation of gene expression of the genes co-expressed with the *SLC6A3* gene in the striatum.

### Comparison between Polygenic risk scores and ePGS

To compare the results obtained with the striatum *SLC6A3* ePGS, we generated traditional polygenic risk scores (PRS) using our accelerated pipeline (https://github.com/MeaneyLab/PRSoS)^57^. A traditional PRS is a cumulative score calculated based on a relevant GWAS that represents a risk for a certain health outcome or trait^58^. The sum of the allele count weighted by the effect size across all SNPs in GWAS at a specified threshold was used to calculate type 2 diabetes^59^ and major depression disorder^60^ PRSs. The number of SNPs included was defined based on the number of SNPs present in our striatum *SLC6A3* ePGS calculated in the discovery cohort. For MDD PRS we used the GWAS results that were obtained without UK Biobank or 23andMe subjects.

### Functional enrichment analysis

Enrichment analysis was performed using MetaCore® software from Clarivate Analytics (https://portal.genego.com) to characterize the putative biological functions associated with the striatal *SLC6A3* co-expression gene network. Genes that comprise the striatal *SLC6A3* ePGS were used in the analysis and the whole genome was used as a background. The significance was considered for the false discovery rate (FDR) adjusted p-value <0.05. To investigate network centrality measures, co-expression patterns were mined from geneMANIA^61^.The gene interactions were then visualized using the Cytoscape® software^62^. The nodes are the elements of a network (genes) and edges are the connection between these elements. Bottleneck genes are defined as those having a high betweenness (the extent to which genes act as ‘bridges’ between other genes in a network), hub genes are defined as those with a high degree (genes with more connections to other genes). To analyze the topological properties associated with this gene network, the CentiScaPe app in Cytoscape® was used to calculate the degree and betweenness of each gene. We used this information to define the “hub genes” within the network, characterized as nodes with degrees higher than +1SD above the mean; and the “bottlenecks” characterized as nodes with betweenness higher than +1SD above the mean. A gene that is both bottleneck and hub was considered as a central node of the network^63^. We also mined protein-protein (PPI) network interactions using the STRING database (https://string-db.org)^64^ and the striatum *SLC6A3* ePGS genes, with the objective to query the physical interactions of the genes that compose our genetic score. Although we mapped the mice co-expressed gene list to human orthologs, not necessarily the co-expression features would be recapitulated in humans. In order to confirm if the genes that comprised the striatal *SLC6A3* ePGS are co-expressed in humans and to analyze their patterns of co-expression during different life periods in humans, we used the gene expression data from human postmortem samples from the BrainSpan database^65^ (see *Supplementary information***, Supplementary Methods**).

### Outcome measures

#### Adult cohort

Psychiatric disorder diagnosis was defined based on the primary or secondary diagnosis of a mental, mood, schizophrenia and neurotic disorders according to participants hospital inpatient records, coded according to the International Classification of Diseases version 10 (ICD-10)^66^ (UK Biobank field 41270; ICD10 codes: F10-F19 Mental and behavioural disorders due to psychoactive substance use, F20-F29 Schizophrenia, schizotypal and delusional disorders, F30-F39 Mood [affective] disorders, F40-F48 Neurotic, stress-related and somatoform disorders). Cardiometabolic disorders diagnosis was defined by the ICD-10 codes from chapter IV Endocrine, nutritional and metabolic diseases and chapter IX Diseases of the circulatory system (UK Biobank fields: 41270; ICD10 codes: E11-Non-insulin-dependent diabetes, I70-Atherosclerosis, I63-Cerebral infarction, I20-I25 Ischaemic heart diseases). The presence of at least one mental disorder diagnosis and at least one cardiometabolic diagnosis was considered a comorbidity case. Comorbidity variable was coded as a binary variable (1 = “yes” or 0 = “no”). T1 structural brain MRI pre-processed imaging data were generated by an image-processing pipeline developed and run on behalf of the UK Biobank^67^ (*Supplementary information***, Supplementary Methods**).

#### Adolescent cohort

No diagnoses for the psychiatric and cardiometabolic disorders noted above were available in ALSPAC. As recommended by the American Academy of Pediatrics (AAP)^68^, we defined disease risk in adolescents using continuous measures of Total difficulties score measured by the Strengths and Difficulties Questionnaire, depression and anxiety scores measured by Computerized Interview Schedule – Revised (CIS-R), Homeostatic Model Assessment of Insulin Resistance (HOMA-IR), and waist circumference (cm) (*Supplementary information,* **Supplementary Methods**, **Supplementary Table 2**, **Supplementary** Figure 3). We then characterized two groups of children: low and high cardiometabolic and psychiatric comorbidity risk (see Statistical Analysis).

### Statistical Analysis

Statistical analysis were performed using R^69^. For the descriptive statistics, the ePGS groups were defined by median split, and a comparison between low and high ePGS groups was done using Student t test for continuous variables and a chi-square test for categorical variables (**Table 1** and **Table 2**). Significance levels for all tests were set at p < 0.05.

We performed cluster analysis using the mclust package to construct the comorbidity risk variable in ALSPAC adolescent sample ^70^. This algorithm applies a model-based classification and density estimation of the z-standardized variables based on finite Gaussian mixture modelling. The method assumes that predictors can be explained by an underlying latent categorical variable (cluster) that represents distinct profiles within the sample, both in a qualitative and quantitative manner. We defined a priori a cluster size solution of two (lower and higher risk for comorbidity). All predictors were z-transformed and adjusted for sex prior to entering the clustering procedure. Regression analysis was carried out to demonstrate the difference between the two clusters in the means for each variable used in the cluster analysis (*Supplementary information***, Supplementary Table 2**). The resulting cluster membership, which represented comorbidity risk, was coded as a binary variable (1= “yes comorbidity” or 0= “no comorbidity”).

The gene by environment (G-E) interaction effect on binary outcomes was explored by logistic regression analysis. Birth weight as a continuous variable was used as a proxy for early life environment exposure in UK Biobank (variable ID20022) and ALSPAC. Early life adversity (E), striatal *SLC6A3* ePGS (G) and the interaction term between them were included in the model as main predictors for both cohorts. UK Biobank analyses were also adjusted by sex, age, the first forty genetic principal components, genotyping array, and assessment center, and ALSPAC analyses were adjusted by sex and the first ten genetic principal components. In case of a significant gene by environment (G-E) interaction effect, post hoc simple slope analysis was performed to investigate how the environment effect varies as a function of the genetic background^71^. The directionality of the G-E effect was explored in the UK Biobank, our discovery cohort, using a two-tailed P-value threshold. The directionality of the G-E effect on ALSPAC was anticipated based on the finding from UK Biobank, thus a one-tailed P-value threshold was considered.

The relation between early life adversity, ePGS and gray matter density in UK Biobank was analyzed in a multivariate parallel independent component analysis (pICA). This analysis was applied to identify the effect of early life adversity on the relation between two different data modalities (genetic and gray matter density) in a data-driven manner^72^. This analysis separately estimates the maximum independent components within each data modality while also maximizing the association between modalities using an entropy term based on information theory^72^. Each SNP that composes the striatal *SLC6A3* ePGS weighted by striatal GTEx data (genotype * GTEx striatum gene expression slope for each SNP) and whole brain voxel based gray matter density were used in the analysis. Weighted SNPs were adjusted for the genetic principal components (ancestry). The subjects were split in two groups according to the birth weight (low birth weight group: subjects with birth weight <= 2.5kg, n = 953) and a randomly selected group of non-low birth weight individuals (subjects with birth weight > 2.5kg, n = 953, please see https://www.who.int/data/nutrition/nlis/info/low-birth-weight), since there was a large discrepancy between cases and controls sample size within the subsample of individuals with T1 structural brain MRI available. Comparison between low birth weight and randomly selected non-low birth weight individuals on main descriptive variables can be seen on Supplementary information (*Supplementary information***, Supplementary Table 5**). Comparison between the randomly selected group and the full sample of non-low birth weight individuals with MRI available can be seen on Supplementary information (*Supplementary information***, Supplementary Table 6**). T1 structural brain MRI pre-processed images were adjusted by age and sex (See *Supplementary information***, Supplementary Methods)**. The Fusion ICA Toolbox (http://mialab.mrn.org/software/fit/) within MATLAB® R2019 was used to run the analysis. The number of independent components was estimated using minimum description length criteria^72^ for the MRI modality and SNP dimensionality inside the toolbox for the genetic modality. Components for both modalities were converted to z-scores and a threshold at |Z| > 2.5 was used to identify significant brain regions and SNPs that contributed the most for the component overall pattern^72^. Loading coefficients, which describe the presence of the identified component across subjects^72^, were extracted for each component, modality, and subject. The mean subject-specific loading coefficients of these components from low birth weight and non-low birth weight groups were compared using Student’s t-test. Talairach coordinates were used to identify the anatomical classification of brain areas included in the identified MRI component^73^. The significant SNPs (|Z| > 2.5) from the identified genetic component were analyzed using MetaCore®, to identify associated gene ontology processes terms (See Figure 3A for graphical representation of pICA analysis).

## Results

### Characteristics of the striatal *SLC6A3* gene network

We developed a polygenic score to explore the genetic moderation of early life adversity on psychiatric and cardiometabolic comorbidities focusing on a specific gene network (Figure 1A **and 1B**). We first used brain region-specific RNA sequencing data from mice available at GeneNetwork (http://genenetwork.org/)^49^ to identify genes co-expressed with the *SLC6A3* gene in the striatum. These genes were then converted to human orthologs (**Supplementary Table 1** and Figure 1B). This list was used to inform the calculation of the expression-based polygenic score (Striatum *SLC6A3* ePGS) in UK Biobank and ALSPAC participants as described in the Methods.

To investigate if mouse-generated *SLC6A3* gene network was co-expressed in humans, we queried the gene co-expression patterns of the striatum *SLC6A3* gene network throughout human development using gene expression data from human postmortem samples^65^. A high co-expression was expected in childhood/adolescence, as the striatum *SLC6A3* gene network was enriched for genes overexpressed in this period of life (see Figure 1C and *Supplementary information***, Supplementary Methods**). Prominent gene co-expression clusters were also seen in adults (Figure 1C). These findings confirm that the striatal *SLC6A3* gene network, originally from murine data, is also co-expressed in humans, and that co-expression is observed at different ages. When visualizing and exploring the network properties (Figure 1D), we observed that the central gene (hub) and the hub-bottleneck genes are related to ribosomal structure. Among the bottleneck genes, which are important connectors between groups of genes, we observed HNRNPA1, which is involved in the packaging of pre-mRNA into particles and transportation from the nucleus to the cytoplasm, as well as splicing. We also observed SDC3 gene, which plays a role in cell shape organization and has been associated with obesity^74^. Protein-protein interactions of the striatum *SLC6A3* co-expression network mined from STRING revealed that the network has significantly more interactions than expected by chance (P<1.0e-16), suggesting that a significant number of the genes in this co-expression network also have physical interactions at the protein level.

The main gene ontology processes terms associated with the network (*Supplementary information***, Supplementary Figure S4)** include: insulin signaling and response terms (Insulin receptor signaling pathway via phosphatidylinositol 3-kinase; Insulin receptor signaling pathway; Cellular response to insulin stimulus; Response to insulin), ribosome production related terms (Ribosome biogenesis; Ribosomal large subunit biogenesis), dopamine receptor signaling pathway (Adenylate cyclase-activating dopamine receptor signaling pathway) and inflammatory response related terms (Regulation of cytokine production involved in inflammatory response; Negative regulation of cytokine production involved in inflammatory response).

### Striatum *SLC6A3* ePGS moderates the association between birth weight and the risk for psychiatric and cardiometabolic comorbidities in adults

Lower birth weight was associated with the presence of comorbidities in UK Biobank (b= -0.206, Odds ratio (OR)= 0.814, 95% confidence interval (95% CI): 0.781 – 0.847, P <0.001). However, in ALSPAC this association was not significant (b= -0.091, OR= 0.913, 95% CI:0.680 – 1.228, P = 0.548).

For the UK Biobank there was no significant main association of the ePGS with comorbidity (b= 0.006, OR= 1.006, 95% CI: 0.977 – 1.036, P=0.678). In contrast, and consistent with our anticipated hypothesis, there was a significant interaction effect between the striatum *SLC6A3* ePGS and birth weight on the presence of psychiatric and cardiometabolic comorbidities in UK Biobank adults (b= 0.042, OR = 1.043, 95% CI: 1.001 – 1.086, P = 0.044). The risk for comorbidity increased as birth weight decreased, especially at lower ePGS values (Low ePGS: b = -0.247, OR = 0.781, P< 0.001, 95% CI 0.738 – 0.826; High ePGS: b = -0.166, OR = 0.847, P< 0.001, 95% CI 0.800 – 0.897).

(Figure 2A). (As we considered birth weight as a continuous variable ranging from low to high values and comorbidity as a dichotomous variable, with the presence of comorbidity computed as 1 and absence as 0, the odds ratio represents the negative association between birth weight and the probability of having comorbidity. Results are presented from the perspective of low birth weight).

**Figure 2.**
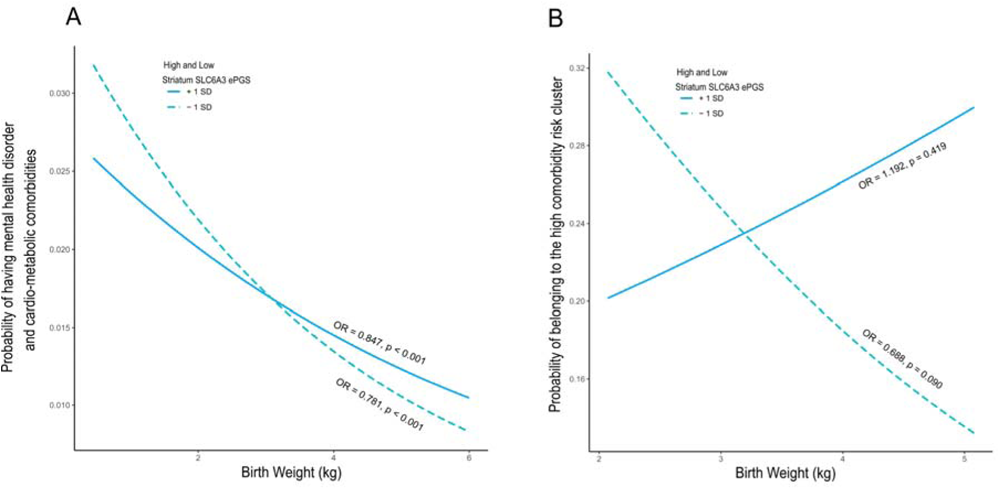
Striatum *SLC6A3* ePGS moderates the effect of early life adversity on the risk for mental health disorder and cardiometabolic comorbidity. Probability of having comorbidity in individuals with high and low striatum *SLC6A3* ePGS as a function of birth weight. A, UK Biobank cohort, N=225,972. The risk for comorbidity increases as birth weight decreases, especially at lower ePGS values (Low ePGS: b = - 0.247, OR = 0.781, P< 0.001, 95% CI 0.738 – 0.826; High ePGS: b = -0.166, OR = 0.847, P< 0.001, 95% CI 0.800 – 0.897). B, ALSPAC cohort, N= 1,188. The risk for comorbidity increases as birth weight decreases, especially at lower ePGS values (Low ePGS: b = -0.373, OR = 0.688, 95% CI 0.447 – 1.066, P=0.090; High ePGS: b = 0.176, OR = 1.192, P = 0.419, 95% CI 0.779 – 1.825).

In ALSPAC adolescents the G-E model revealed a significant interaction effect between the striatum *SLC6A3* ePGS and birth weight on the probability of belonging to the high comorbidity risk cluster (b = 0.271, OR= 1.311, 95% CI: 1.015 – Inf, P = 0.041, n=1,188). The risk for of belonging to the high comorbidity risk cluster increased as birth weight decreased, especially at lower ePGS values (Low ePGS: b = -0.373, OR = 0.688, 95% CI 0.447 – 1.066, P=0.090; High ePGS: b = 0.176, OR = 1.192, P = 0.419, 95% CI 0.779 – 1.825). (Figure 2B). Similar to the findings in adults, there was no significant main effect association of the striatum *SLC6A3* ePGS on the comorbidity risk (b= 0.086, OR= 1.090, 95% CI: 0.957 – 1.240, P= 0.195). These results indicate a developmental trajectory, in which early indicators of risk to develop psychiatric and metabolic comorbidities in adulthood can be seen in adolescents as a function of the interaction of the striatum *SLC6A3* co-expression gene network and birth weight.

To benchmark our method against the classical polygenic risk score derived from a GWAS, we performed the same G-E interaction analysis using birth weight and PRSs based on GWAS for major depressive disorder^60^ and type 2 diabetes^59^. These are phenotypes related to our main outcome, psychiatric and cardiometabolic comorbidity.

We found significant main effects of Type 2 diabetes and MDD PRSs on the comorbidity outcome in UK Biobank, but not in ALSPAC (*Supplementary information***, Supplementary Table 3**) and no significant G-E interaction on comorbidity using these PRSs in the UK Biobank or ALSPAC (*Supplementary information***, Supplementary Table 4**).

### SNPs from the striatum *SLC6A3* ePGS are related to gray matter variations in the frontal cortex

We then explored the neuroanatomo-functional relevance of the relation between the striatal *SLC6A3* gene network and early adversity. Functional refers to the variation in gene expression represented by the weight attributed to the SNPs that compose the striatum *SLC6A3* ePGS. We used a multivariate parallel independent component analysis (pICA)^72^ (Figure 3A and *Supplementary information***, Supplementary Methods**) and investigated correlations between the SNPs from the striatum *SLC6A3* ePGS and voxel-based gray matter density in UK Biobank participants from low birth weight and non-low birth weight groups. This analysis identifies independent components within each data modality separately (SNPs and MRI) while also maximizing the association between these two modalities. The estimated number of components for the MRI modality was 28 and for the genetic modality was 34. Only the most significantly linked pair of components that resulted from the multivariate analysis with higher correlation index value was selected to be further explored: the pair combining the genetic component 13 and MRI component 18 (r=-0.201, p=6.779e^-19^). A statistically significant difference between birth weight groups was observed for both the genetic component 13 (t=2,214, p=0.026) as well as the MRI component 18 (t=-3,318, p<0.001). These differences between the adversity groups suggest that the relations between data pattern variations (i.e., the relationships between SNPs and gray matter) within this pair of components are significantly different between the two birth weight groups. We then explored the content of genetic component 13 and MRI component 18. The subset of significant SNPs within component 13 is related to variations in gray matter density in the frontal cortex, including the prefrontal cortex, and also more specifically the orbitofrontal cortex, part of the prefrontal cortex, cingulate cortex and temporal cortex (Figure 3B). Enrichment analysis of this subset of significant SNPs (*Supplementary information* **Supplementary Table 7**) using MetaCore® (FDR<0.05) showed that the most significant gene ontology enrichment terms are related to regulation of dendrite development, regulation of neuron remodeling, positive regulation of nervous system development, pyruvate biosynthetic process, ATP metabolic process and response to epinephrine (Figure 3C).

**Figure 3.**
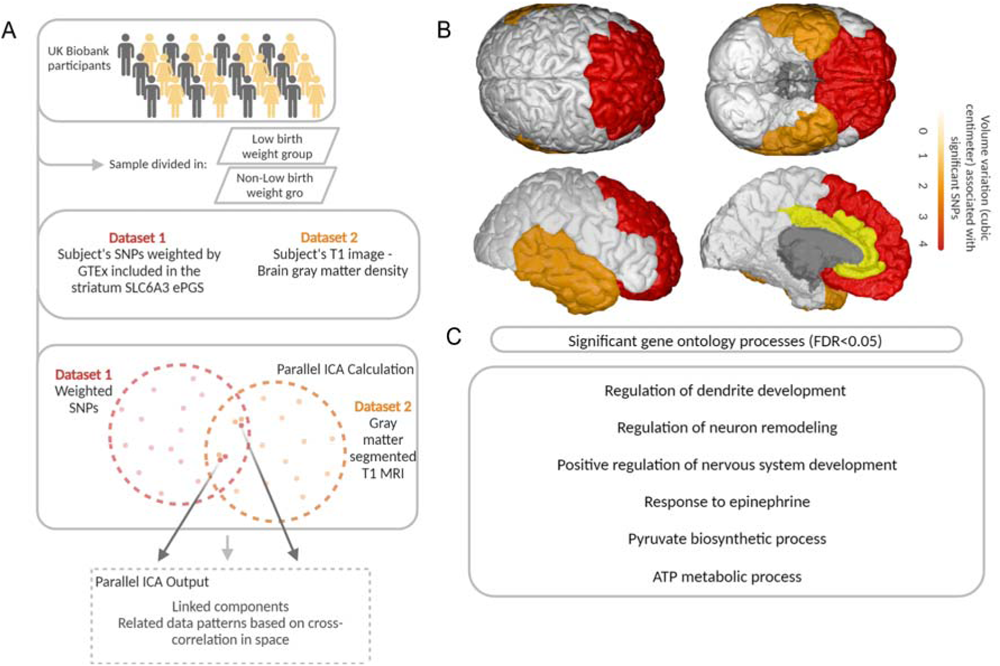
Parallel ICA analysis. A, schematic representation of parallel ICA method. Two different data modalities (SNPs and voxel-based gray matter) were used to establish anatomical-functional correlations between the striatum *SLC6A3* ePGS and brain features from UK Biobank participants (N=11,167). Participants were separated into low birth weight and normal birth weight groups. The analysis estimates the maximum independent components within each data modality separately while also maximizing the association between modalities using an entropy term based on information theory. B, significant brain regions associated with SNPs from the striatum *SLC6A3* were frontal cortex, including the orbitofrontal and prefrontal cortex, cingulate cortex and temporal cortex. Color scheme represents the amount of volume variation (cubic centimeter) significantly associated with the subset of SNPs. C, summary of significant gene ontology processes related to SNPs from the striatum *SLC6A3* ePGS associated with gray matter.

## Discussion

Our study suggests that being born with lower birth weight increases the risk for later comorbidities between cardiometabolic and psychiatric conditions in adulthood. In fact, being born at low birth weight, which reflects prenatal adversity^13^, independently associates with increased risk for developing both cardiometabolic^15–18^ and psychiatric disorders^19–24^ corroborating our findings. Our functional genomics approach provides evidence for the striatal *SLC6A3* co-expression gene network as a salient mechanism moderating this association. This finding is aligned with the critical role of the dopaminergic system in environmental responsivity^42,75^.

Although lower birth weight is associated with an increased risk for co-morbidity in both ePGS groups in the UK Biobank, low ePGS participants have significantly more risk than high ePGS individuals. In the low ePGS group in adolescence, there is a suggestion of increased risk of being part of a high comorbidity risk as birth weight decreases (p=0.09). Although the simple slope for the high ePGS group in adolescence shows a positive inclination between birth weight and risk for comorbidity, this slope is not significant therefore the risk for comorbidity does not vary according to birth weight in the high ePGS group. No information on gestational age was available in the UK Biobank cohort, to maintain consistency, birth weight as a continuous variable and not corrected for gestational age was used in both cohorts. The lack of information about gestational age in our study may be especially affecting the adolescent analysis and this may potentially explain why the simple slope for the low ePRS group does not reach statistical significance, although the interaction between ePRS and birth weight is statistically significant in ALSPAC.

Our enrichment analysis showed that the striatal *SLC6A3* gene network is co-expressed in humans across childhood/adolescence and adulthood (Figure 1C), which is aligned with our interaction between striatum *SLC6A3* ePGS and birth weight been observed both in adolescents and in adults. Our results therefore demonstrate that striatum *SLC6A3* ePGS is able to detect individual differences in response to early adversity at both ages. Based on the comparisons observed in this study, only the striatum *SLC6A3* ePGS was capable of capturing gene by environment interaction effects, while different PRSs did not significantly interact with early life adversity to predict the main outcome. GWAS-derived PRS reflect main genetic effects and thus are unlikely to capture individual differences in response to environmental variation. Indeed we found significant main effects of the PRSs of Major depressive disorder^60^ and Type 2 diabetes ^59^ on comorbidity in UK Biobank. Overall, these results align with the well-known capacity of PRS to detect main genetic effects, as well as demonstrate the ability of our ePGS technique in identifying responsivity to environmental change as compared to traditional GWAS-based PRS^76^. PRS main effects were not observed in adolescents from ALSPAC probably due to the specificity of the GWAS to the features of the original discovery sample; the majority of GWASs are generated based on adult samples^77^, thus limiting the extrapolation of the effects in different ages.

We also identified putative biological mechanisms underlying the moderating effect of the striatum *SLC6A3* ePGS on the association between early adversity and cardiometabolic – psychiatric comorbidities (Figure 1D and **Supplementary** Figure 4). Central genes of the striatum *SLC6A3* network are related to ribosomal structure and the entire gene network is significantly enriched for ribosome production related terms (Ribosome biogenesis; Ribosomal large subunit biogenesis) as seen in our gene-ontology analysis. Ribosomes functioning is highly related to cell growth and proliferation and protein synthesis in all cells. The ribosome biogenesis is a critical process to form mature ribosomes^78^. Dysfunction of ribosomal gene expression has been seen in animals’ models of depression^79^ and the use of anti-ribosomal P antibodies, that targets phosphorylated protein (P) components of ribosomes, was able to induce depression like behavior in mice^80^. Interestingly, the same antibody is used to detect systemic lupus erythematosus in humans^80^. This autoimmune disease is related to high levels of systemic inflammation. Our *SLC6A3* gene network is significantly enriched for inflammatory response related terms, especially cytokine production. One of the central genes of our network, *GNB1*, has been implicated in regulation of inflammasomes, multiple proteins complexes responsible for activation of inflammatory responses^81^. In fact, many patients with major depressive disorder have elevated levels of inflammatory cytokines ^82^ and there is evidence linking cardiometabolic syndrome to higher levels of circulating cytokines^83^. Not surprisingly, our *SLC6A3* gene network is also significantly enriched for dopamine receptor signaling pathway. *HNRNPA1*, a central node of our network, is involved in mRNA transport and synthesis^84^. mRNA axonal transport and protein synthesis at the terminal is an important mechanism for regulation of neurotransmitter synthesis and reuptake^85^. Although *SLC6A3* gene expression occurs at the level of the ventral tegmental area, having *HNRNPA1* as a central node of our network could explain the presence of *SLC6A3* mRNA at the striatal terminal, consistent with numerous human post-mortem studies^86–89^. *SLC6A3* mRNA transport to terminals may be a key mechanistic feature of our *SLC6A3* striatum gene network. It has been shown that *SLC6A3* protein vesicular traffic has a limited contribution to *SLC6A3* concentration in synapses^90^, hence other forms of regulation of *SLC6A3* availability in terminals – for instance via mRNA axonal transport and terminal protein synthesis – are likely in place, in agreement to our findings.

Another important gene in the network is the *SDC3*, that may play a role in cell shape organization and has been associated with obesity^74^. Obesity is related to increased risk for cardiovascular disease^91^, high levels of inflammation and insulin resistance^92^. Our striatum *SLC6A3* gene network is significantly enriched for insulin signaling and response related terms. Being born with low birth weight is associated with insulin resistance in children and adolescents^93^ and insulin resistance is a risk factor for cardiometabolic and brain-based disorders, including type II diabetes, cardiovascular disease, Alzheimer disease, and major depressive disorder^94,95^. Metformin, a medication to treat insulin resistance, has shown beneficial psychotropic effects in psychiatric conditions^96,97^. Evidence shows that insulin has a role in modulating mesocorticolimbic DA neurotransmission through different mechanisms, one of which is increasing DA reuptake by activating the phosphatidylinositol (PI) 3-kinase^36,98^. Insulin also reduces DA release in rodent nucleus accumbens and medial prefrontal cortex slices^99^. Our significant gene by environment results, using birth weight as our environmental proxy, corroborate with the literature showing elevated risk for developing psychiatric and cardiometabolic disorders among individuals born with low birth weight. Our genetic enrichment analysis results indicate that insulin signaling disturbances may be a potential mechanism involved in the interaction effect between birth weight and the striatum *SLC6A3* co-expression gene network on the risk for cardiometabolic and psychiatric comorbidities. This is aligned with many other studies suggesting that altered insulin function is an important mechanism linking early adversity to later disease^100,101^.

The subset of SNPs from the striatum *SLC6A3* ePGS that is related to gray matter density variations in our neuroanatomical-functional correlation analysis is associated with regulation of dendrite morphogenesis, neuron remodeling and positive regulation of nervous system development. These are important processes linked to the prolonged maturation of the mesocorticolimbic dopamine system during the life-course^102^, which makes striatal dopaminergic axons especially vulnerable to environmental effects during development^103^. According to these findings, we recently showed that both rodent poor fetal growth and insulin treatment affect the expression of the Netrin-1/DCC axonal guidance cue system, which is involved in the maturation of the mesocorticolimbic DA circuitry^104^. Pyruvate biosynthetic process and ATP metabolic process also emerged as significant enrichment terms for the subset of significant SNPs related to gray matter density. Both processes have connections with insulin secretion: regulation of insulin secretion in pancreatic β cells is modulated by ATP synthesis and release in mitochondria^105^ and by pyruvate transport through mitochondrial pyruvate carriers^106^. The frontal, prefrontal and orbitofrontal cortices were related to the significant subset of SNPs identified by the pICA analysis. This is aligned with evidence demonstrating that resting state functional connectivity between the orbitofrontal cortex and dorsolateral prefrontal cortex is altered in human individuals born small for gestational age, at different ages during development^107^.

The cingulate and temporal cortices also emerged as a significant brain regions in the pICA analysis. The anterior cingulate has been implicated in affective abnormalities in mood disorders and volume reduction in patients with major depressive disorder^108^. Abnormal posterior cingulate functional connectivity has also been reported in major depression^109^. Temporal lobe alterations are related to insulin resistance pathophysiology in different imaging modalities^110^. Interestingly, the relationship between genetic and MRI components was significantly different between the two early life adversity groups, suggesting that the biological mechanisms represented by the genetic component and the brain regions highlighted by the MRI component are relevant for the effects of early adversity on adult disease.

Taken together, the evidence suggests that ribosomal function, inflammation, and insulin modulation of dopamine function may be underlying mechanisms by which the striatum *SLC6A3* gene network moderates the risk for developing psychiatric and cardiometabolic comorbidities in response to early life adversity. These mechanisms might be especially important in brain areas involving the prefrontal and orbitofrontal cortices, cingulate and temporal cortices.

Our study is limited by the fact that ePGS does not consider intronic regions, potentially ignoring other important regulatory elements. Moreover, our developmental results are based on cross sectional studies, and further longitudinal data are needed to better describe this trajectory.

In sum, we observed that the association between environmental and genetic factors can place individuals at risk for adult comorbid chronic conditions from an early age, and that a striatal dopamine transporter gene network expression has a central role in moderating the association of the early environment with the risk for these diseases. These findings open opportunities for the exploration of the understudied field of precision prevention in pediatrics, and the potential design of more effective interventions and primary care strategies.

## Data Availability

For UK Biobank, data can be purchased; the study website contains details of all the data that is available at https://biobank.ndph.ox.ac.uk/showcase/.
For ALSPAC, data can be purchased; the study website contains details of all the data that is available through a fully searchable data dictionary and variable search tool at http://www.bristol.ac.uk/alspac/researchers/our-data/.

## Acknowledgments

We are extremely grateful to all the families who took part in this study, the midwives for their help in recruiting them, and the whole ALSPAC and UK Biobank teams, which includes interviewers, computer and laboratory technicians, clerical workers, research scientists, volunteers, managers, receptionists and nurses. The UK Medical Research Council and Wellcome (Grant ref: 217065/Z/19/Z) and the University of Bristol provide core support for ALSPAC. This publication is the work of the authors and Patricia Pelufo Silveira and Barbara Barth will serve as guarantors for the contents of this paper. A comprehensive list of grants funding is available on the ALSPAC website (http://www.bristol.ac.uk/alspac/external/documents/grant-acknowledgements.pdf). This research using ALSPAC was specifically funded by the Wellcome Trust and MRC (Grant ref: 076467/Z/05/Z) and Wellcome Trust (Grant ref: 08426812/Z/07/Z). This project was supported by The JPB Foundation through a grant to The JPB Research Network on Toxic Stress: A Project of the Center on the Developing Child at Harvard University. This work was also funded by the Fonds de recherche du Québec – Santé and Canadian Institutes of Health Research (CIHR, PJT-166066, PI Silveira PP).

