## Supplementary material for "Striatal dopamine gene network moderates the effect of early adversity on the risk for adult psychiatric and cardiometabolic comorbidity"

**Supplementary Methods**

**Participants**

*UK Biobank – Adult cohort*: UK Biobank contains information on participants' lifestyle and health data at baseline or follow-up, which were collected through questionnaires, physical measurements, and biological samples. For the purpose of this project, only unrelated subjects were considered in the analysis. Exclusion criteria were 1) participants who withdrew their consent from the study, 2) no genotyping data, 3) related participants (genetic kinship to other participants > 0.04), 4) inconsistencies in genetic and reported sex and 5) outliers for heterozygosity. All subjects selected for the analysis satisfied the following criteria: (1) have genotyping data available and (2) have both the diagnosis outcome and birth weight available. Detailed description of sample selection process can be found in **Supplementary Figure 1**.

*ALSPAC – Adolescent cohort*: Data were collected during clinic visits or with postal questionnaires. Please note that the study website contains details of all the data that is available through a fully searchable data dictionary and variable search tool at <http://www.bristol.ac.uk/alspac/researchers/our-data/>. The following inclusion criteria were applied: unrelated individuals, gestational age between 37 and 42 weeks inclusively, maternal age at delivery >= 18 years old, birth weight of at least 2000 grams, singleton pregnancies. Detailed description of sample selection process can be found in **Supplementary Figure 2**. The participant attrition block scheme differs slightly in originally projected ALSPAC participant numbers as we are using an earlier data file from 2019 to complete these analyses.

**Genotyping**

*UK Biobank – Adult cohort*: Blood samples from the UK Biobank were genotyped at the Affymetrix Research Services Laboratory in Santa Clara, California, USA. Genotyping was conducted using a bespoke BiLEVE Axiom array for 50,000 participants and the remaining 450,000 participants were genotyped using the Affymetrix UK Biobank Axiom array. The two SNP arrays are very similar with over 95% common marker content. Axiom Array plates were processed on the Affymetrix GeneTitan® Multi-Channel (MC) Instrument. Genotypes were then called from the resulting intensities in batches of ~4,700 samples (~4,800 including the controls) using the Affymetrix Power Tools software and the Affymetrix Best Practices Workflow. Individuals with the same genotype at any given SNP will cluster together in a two-dimensional intensity space (one dimension for each targeted allele). For the interim data release, Affymetrix performed further rounds of genotype calling using algorithms customized for the UK Biobank project. These algorithms targeted very rare SNPs with 6 or fewer minor alleles in a batch, and a subset of SNPs for which the generic calling algorithm did not perform optimally. After genotype calling, Affymetrix performed quality control in each batch separately, to exclude SNPs with poor cluster properties. If a SNP did not meet the Affymetrix prescribed QC thresholds in a given batch, it was set to missing in all individuals from that batch. Hardy-Weinberg equilibrium was performed for each batch. Affymetrix also checked sample quality (such as DNA concentration) and genotype calls were provided only for samples with sufficient DNA metrics. For SNP-based QC metrics, only individuals with similar ancestry and the population structure were characterized by computing principal components using only UK Biobank individuals. The array also includes coding variants across a range of minor allele frequencies (MAFs), including rare markers (<1% MAF); and markers that provide good genome-wide coverage for imputation in European populations in the common (>5%) and low frequency (1–5%) MAF ranges. More information about the genotyping protocol, QC and imputation could be found in ^1^. The population structure of the UK Biobank cohort was evaluated using fastPCA algorithm for principal component analysis^2^. To account for population stratification, the first forty principal components were included in the UK Biobank analysis.

*ALSPAC – Adolescent cohort*: Subjects in ALSPAC cohort were genotyped using the Illumina HumanHap550 quad genome-wide SNP genotyping platform by the Welcome Trust Sanger Institute (Cambridge, UK) and the Laboratory Corporation of America (Burlington, NC, US)^3^. The following quality control procedure was applied: participants with inconsistencies in self-reported and genotyped sex, minimal or extreme heterozygosity, high levels of individual missingness (>3%), and insufficient sample replication (IBD < 0.8) were excluded. SNPs with MAF <1%, call rate <95%, or those not in HWE (p < 5 x 10−7) were removed. Imputation was conducted using Impute v3 and Haplotype Reference Consortium (HRC) imputation reference panel (release 1.1). The resulting data set consisted of 8,365 individuals and 38,898,739 SNPs available for analysis. The population structure of the ALSPAC cohort was described using principal component analysis^4,5^, which was conducted on the genotyped autosomal SNPs with MAF > 5% with the following pruning parameters for linkage disequilibrium: 100-SNP sliding window, an increment of 5 SNPs, and variance inﬂation factor threshold of 1.01. To account for population stratiﬁcation, the ﬁrst ten PCs were included in the analysis. Processing of the genotyping data was done using PLINK 1.9 ^6^(authors: Shaun Purcell, Christopher Chang; www.cog-genomics.org/plink/1.9/).

**Gene expression levels at different developmental stages**

In order to confirm if the genes that composed the striatum SLC6A3 ePGS are co-expressed in humans and investigate their patterns of gene co-expression during different life periods, we used the human postmortem striatal gene expression data from the BrainSpan database^7^. The co-expression patterns were analyzed during two different stages of development: childhood/adolescence (0 to 19 years old, N=7) and adulthood (20 to 40 years old, N=6). The analyses were carried out in R (https://www.r-project.org)^8^ using the heatmaply package^9^.

**Outcome measures**

*UK Biobank – Adult cohort:* To access the presence of psychiatric and cardio-metabolic disorders within the UK Biobank cohort, we utilized diagnostic terms from across all participants hospital inpatient records, coded according to the International Classification of Diseases version 10 (ICD-10)^23^. The presence of at least one cardio-metabolic diagnosis and at least one psychiatric diagnosis was considered a comorbidity case.

**Psychiatric disorder diagnosis definition:**

**Cases:**

From Hospital Episodes Data from UK bodies (English HES Data, Scottish Morbidity Register, Patient Episode Data) (Fields 41270):

Any primary or secondary diagnosis of ICD-10 Codes for:

o ICD10 F10-F19 Mental and behavioural disorders due to psychoactive substance use'

o ICD10 F20-F29 Schizophrenia, schizotypal and delusional disorders

o ICD10 F30-F39 Mood [affective] disorders

o ICD10 F40-F48 Neurotic, stress-related and somatoform disorders

**Controls:**

No primary or secondary diagnosis of ICD-10 codes for:

o ICD10 F10-F19 Mental and behavioural disorders due to psychoactive substance use'

o ICD10 F20-F29 Schizophrenia, schizotypal and delusional disorders

o ICD10 F30-F39 Mood [affective] disorders

o ICD10 F40-F48 Neurotic, stress-related and somatoform disorders

**Cardio-metabolic disorder diagnosis definition:**

**Cases:**

From Hospital Episodes Data from UK bodies (English HES Data, Scottish Morbidity Register, Patient Episode Data) (Fields 41270):

Any primary or secondary diagnosis of ICD-10 Codes for:

o ICD10 E11 Non-insulin-dependent diabetes

o ICD10 I70 Atherosclerosis

o ICD10 I63 Cerebral infarction

o ICD10 I20-I25 Ischaemic heart diseases

**Controls:**

No primary or secondary diagnosis of ICD-10 codes for:

o ICD10 E11 Non-insulin-dependent diabetes

o ICD10 I70 Atherosclerosis

o ICD10 I63 Cerebral infarction

o ICD10 I20-I25 Ischaemic heart diseases

*ALSPAC – Adolescent cohort*: The indicators of risk to develop psychiatric disorders comprised: a) total difficulties score as measured by the Strengths and Difficulties Questionnaire^24^ filled out by primary caregivers of 16.6 year old participants. This instrument evaluates behavioral problems, and a total difficulties score is computed by adding four domains of the scale (emotional symptoms, conduct problems, hyperactivity/inattention and peer relationship problems) that represent negative behaviors; b) depression score and c) anxiety score as measured by the Computerized Interview Schedule – Revised (CIS-R) that establishes the nature and severity of neurotic symptoms^25^, applied to 17.5 year old participants. The indicators of risk to develop metabolic disorders involved: d) insulin resistance as measured by the Homeostatic Model Assessment of Insulin Resistance (HOMA-IR), calculated using plasma fasting glucose (mmol/l) and insulin (pmol/l) levels collected at 17.5 years of age. The calculation followed the updated version of the HOMA-IR index developed by Wallace et al (2004)^26^ and was computed using the HOMA2 calculation tool (http://www.dtu.ox.ac.uk/homacalculator/); and e) waist circumference (cm) measured at 15.5 years of age. Similar to UK Biobank, we created comorbidity risk variable based on the indicators of risk to develop psychiatric and metabolic disorders. Precisely, to construct comorbidity risk variable we performed cluster analysis on five risk indicators: Total difficulties, depression and anxiety scores, HOMA-IR and waist circumference. All predictors were z-transformed prior to entering the clustering procedure and adjusted by sex. We defined a cluster solution of two clusters, representing lower and higher risk for comorbidity. Regression analysis was carried out to demonstrate the difference between the means for each variable included in the cluster analysis (*Supplementary information***, Supplementary Table 2, Supplementary Figure 3**).

*Gray matter density in UK Biobank participants*: T1 structural brain MRI pre-processed imaging data were generated by an image-processing pipeline developed and run on behalf of the UK Biobank^27^. High-resolution T1-structural images for the whole brain were acquired with straight sagittal orientation using a Siemens Skyra 3T running VD13A SP4, with a standard Siemens 32-channel RF receive head coil. The following parameters were used: resolution 1x1x1mm; field-of-view 208x256x256 matrix; 5 minutes duration; 1 mm isotropic resolution using 3D MPRAGE acquisition; in-plane acceleration iPAT=2; prescan-normalize. Full 3D gradient distortion correction (GDC) was applied to the original T1 image and the field of view (FOV) was cut down to reduce the amount of non-brain tissue. Tools used to achieve this include BET (Brain Extraction Tool), FLIRT (FMRIB's Linear Image Registration Tool), and the MNI152 “nonlinear 6th generation” standard-space T1 template. A non-linear registration to MNI152 space was used with FNIRT (FMRIB's Nonlinear Image Registration Tool). Using the inverse of the MNI152 alignment warp, a standard-space brain mask was transformed into the native T1 space and applied to the T1 image to generate a brain-extracted T1. Tissue-type segmentation was applied to T1 weighted images using FSL/FAST (FMRIB's Automated Segmentation Tool). Then, the T1-weighted gray matter images were selected and adjusted for age and sex. For each voxel separately, we applied linear regression analysis to regress the intensity on age and sex and used the residuals in p-ICA analysis.

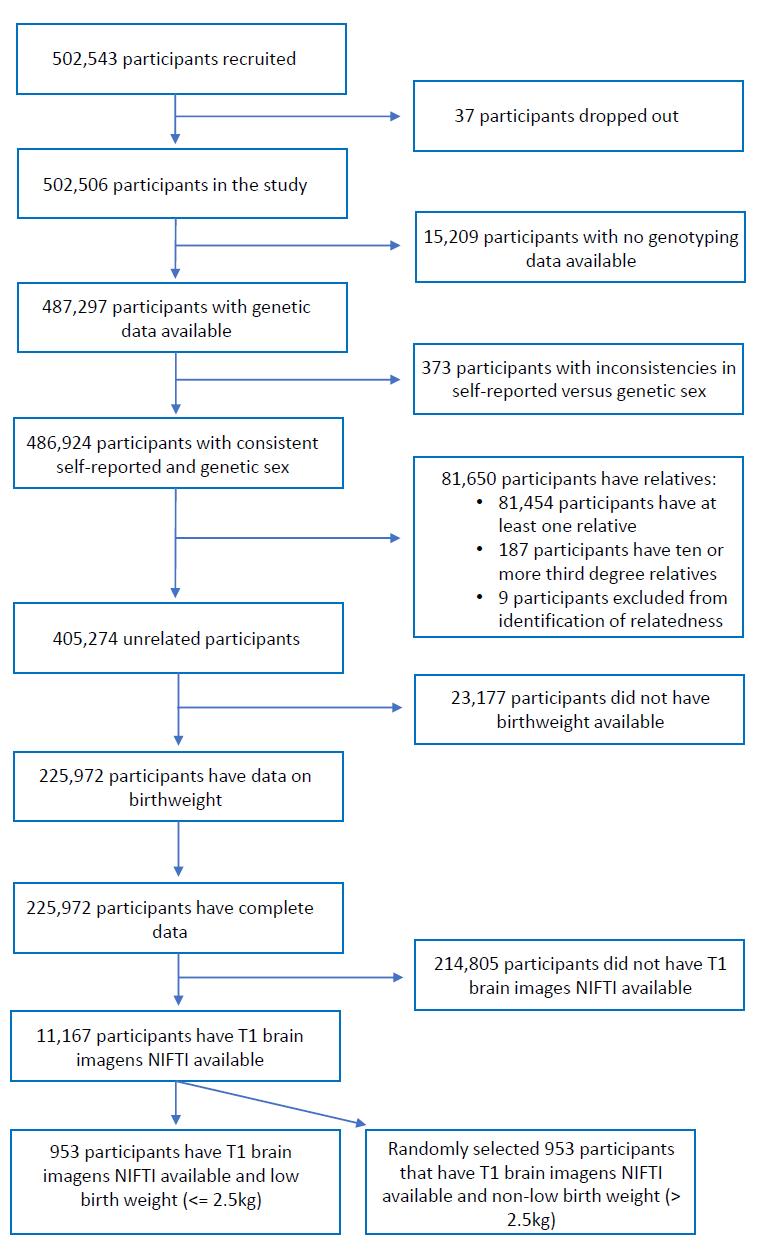

**Supplementary Figure 1**. UK Biobank Sample size block scheme for each inclusion and exclusion criteria applied. Block scheme depicting each step of inclusion and exclusion criteria applied to the original sample (N=502,543) until reaching final sample size for main hypothesis testing (N=225,972) and final sample size for parallel ICA analysis group of low birth weight (N=953) and non-low birth weight (N=953).

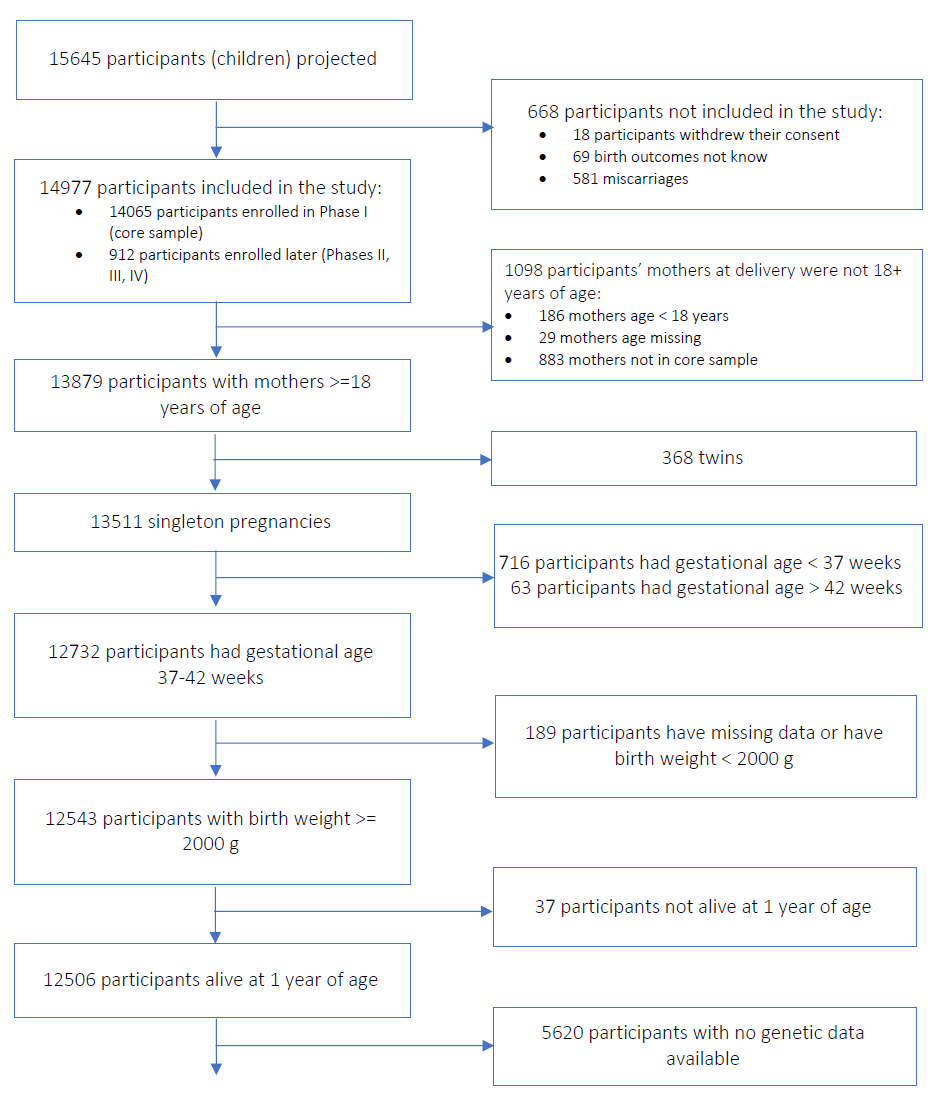

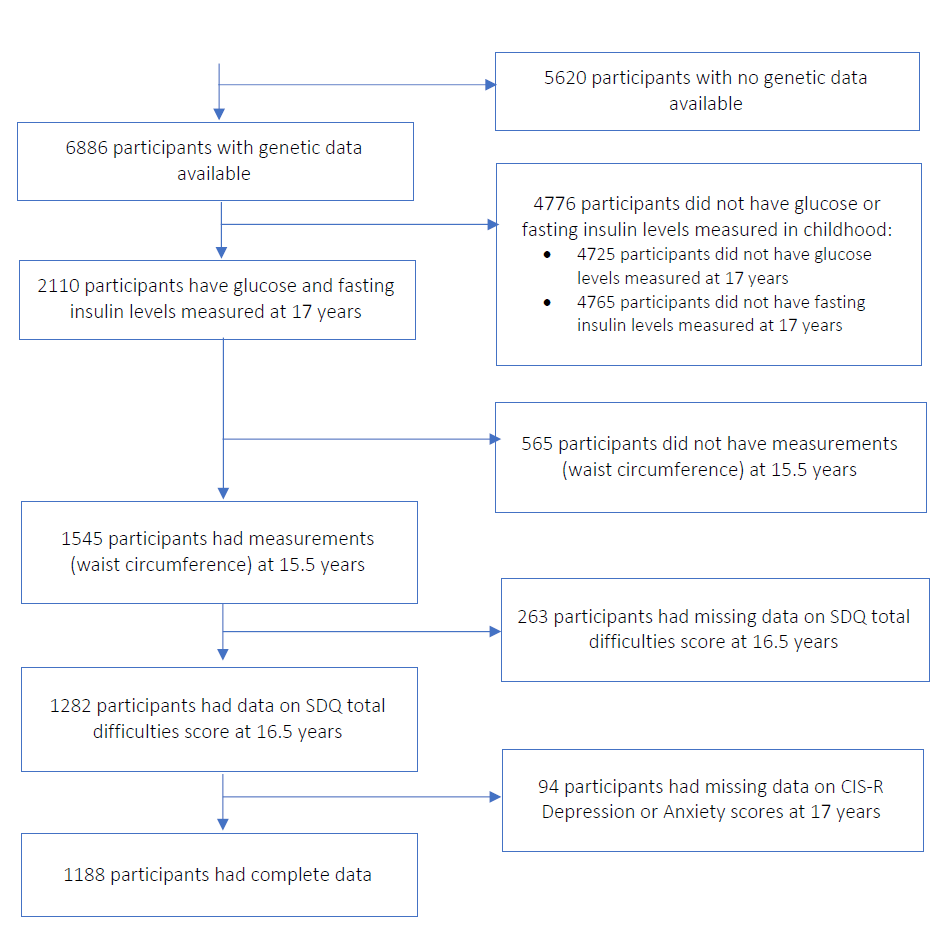

Supplementary Figure 2. ALSPAC Sample size block scheme for each inclusion and exclusion criteria applied Block scheme depicting each step of inclusion and exclusion criteria applied to the original sample (N=15,645) until reaching final sample size for main hypothesis testing (N=1,188).

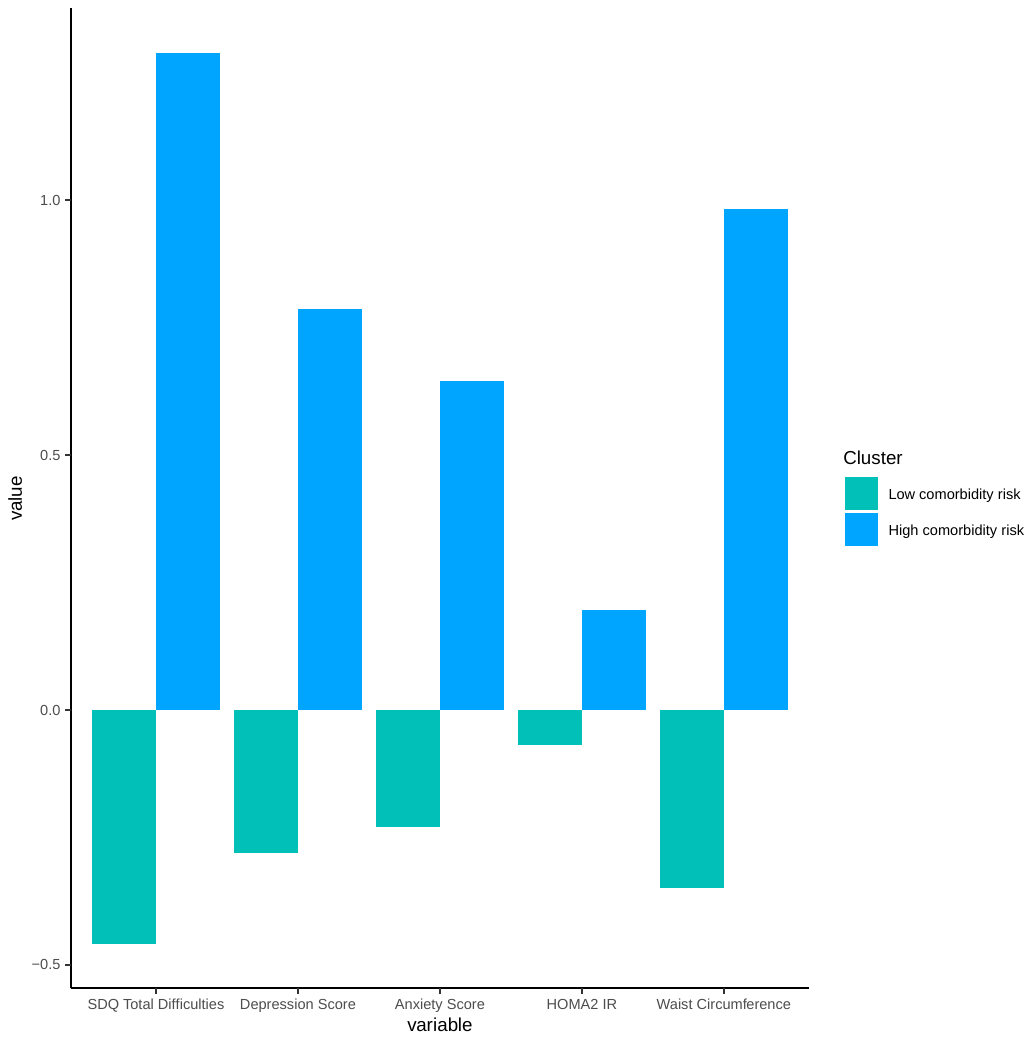

Supplementary Figure 3. Comorbidity risk clusters – ALSPAC. Cluster 1 – Low Comorbidity Risk (N=876) and Cluster 2 – High Comorbidity Risk (N=312) mean z score values for the variables considered in the cluster analysis. Cluster high comorbidity risk represents the psychiatric and metabolic comorbidity risk profile in adolescents. SDQ Total Difficulties is a score from the Strengths & Difficulties Questionnaire (SDQ). Depression and anxiety scores were computed from the Computerized Interview Schedule – Revised (CIS-R). Homeostatic Model Assessment of Insulin Resistance (HOMA-IR) was computed using the HOMA2 calculation tool. Waist circumference was measured in centimeters. Clustering of these variables was performed using mclust package in R.

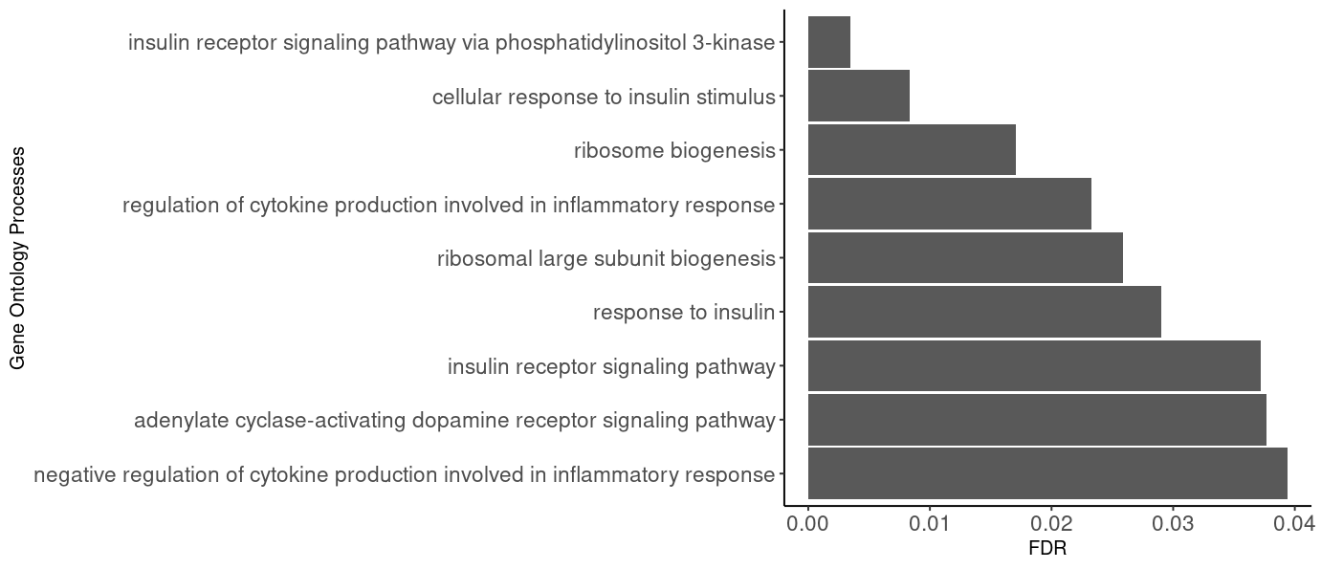

Supplementary Figure 4. Striatal SLC6A3 gene network enrichment analysis. Gene ontology processes related to genes included in striatal SLC6A3 gene network. Enrichment was performed using MetaCore®. The significance was considered for the false discovery rate (FDR) adjusted p-value <0.05.

Supplementary Table 1. Genes co-expressed with the SLC6A3 in mice striatum data, used to calculate striatum *SLC6A3* ePGS genetic score.

| Mouse gene | Mouse Ensembl gene ID | Human gene | Human Ensembl gene ID | Description |
| --- | --- | --- | --- | --- |
| Rnf165 | ENSMUSG00000025427 | RNF165 | ENSG00000141622 | ring finger protein 165 [Source:HGNC Symbol;Acc:HGNC:31696] |
| Rps21 | ENSMUSG00000039001 | RPS21 | ENSG00000171858 | ribosomal protein S21 [Source:HGNC Symbol;Acc:HGNC:10409] |
| Ipmk | ENSMUSG00000060733 | IPMK | ENSG00000151151 | inositol polyphosphate multikinase [Source:HGNC Symbol;Acc:HGNC:20739] |
| Mllt3 | ENSMUSG00000028496 | MLLT3 | ENSG00000171843 | MLLT3 super elongation complex subunit [Source:HGNC Symbol;Acc:HGNC:7136] |
| Hnrnpk | ENSMUSG00000021546 | HNRNPK | ENSG00000165119 | heterogeneous nuclear ribonucleoprotein K [Source:HGNC Symbol;Acc:HGNC:5044] |
| Rsl1d1 | ENSMUSG00000005846 | RSL1D1 | ENSG00000171490 | ribosomal L1 domain containing 1 [Source:HGNC Symbol;Acc:HGNC:24534] |
| Gfra2 | ENSMUSG00000022103 | GFRA2 | ENSG00000168546 | GDNF family receptor alpha 2 [Source:HGNC Symbol;Acc:HGNC:4244] |
| Nptxr | ENSMUSG00000022421 | NPTXR | ENSG00000221890 | neuronal pentraxin receptor [Source:HGNC Symbol;Acc:HGNC:7954] |
| Tgif2 | ENSMUSG00000062175 | TGIF2 | ENSG00000118707 | TGFB induced factor homeobox 2 [Source:HGNC Symbol;Acc:HGNC:15764] |
| Jarid2 | ENSMUSG00000038518 | JARID2 | ENSG00000008083 | jumonji and AT-rich interaction domain containing 2 [Source:HGNC Symbol;Acc:HGNC:6196] |
| Bmf | ENSMUSG00000040093 | BMF | ENSG00000104081 | Bcl2 modifying factor [Source:HGNC Symbol;Acc:HGNC:24132] |
| 6820408C15Rik | ENSMUSG00000032680 | C20orf96 | ENSG00000196476 | chromosome 20 open reading frame 96 [Source:HGNC Symbol;Acc:HGNC:16227] |
| Rpl14 | ENSMUSG00000025794 | RPL14 | ENSG00000188846 | ribosomal protein L14 [Source:HGNC Symbol;Acc:HGNC:10305] |
| Stk32a | ENSMUSG00000039954 | STK32A | ENSG00000169302 | serine/threonine kinase 32A [Source:HGNC Symbol;Acc:HGNC:28317] |
| Cdh10 | ENSMUSG00000022321 | CDH10 | ENSG00000040731 | cadherin 10 [Source:HGNC Symbol;Acc:HGNC:1749] |
| Sdc3 | ENSMUSG00000025743 | SDC3 | ENSG00000162512 | syndecan 3 [Source:HGNC Symbol;Acc:HGNC:10660] |
| Mkrn1 | ENSMUSG00000029922 | MKRN1 | ENSG00000133606 | makorin ring finger protein 1 [Source:HGNC Symbol;Acc:HGNC:7112] |
| Bbc3 | ENSMUSG00000002083 | BBC3 | ENSG00000105327 | BCL2 binding component 3 [Source:HGNC Symbol;Acc:HGNC:17868] |
| Tubb2a | ENSMUSG00000058672 | TUBB2A | ENSG00000137267 | tubulin beta 2A class IIa [Source:HGNC Symbol;Acc:HGNC:12412] |
| Bcl11a | ENSMUSG00000000861 | BCL11A | ENSG00000119866 | BAF chromatin remodeling complex subunit BCL11A [Source:HGNC Symbol;Acc:HGNC:13221] |
| Rpl37a | ENSMUSG00000046330 | RPL37A | ENSG00000197756 | ribosomal protein L37a [Source:HGNC Symbol;Acc:HGNC:10348] |
| Cotl1 | ENSMUSG00000031827 | COTL1 | ENSG00000103187 | coactosin like F-actin binding protein 1 [Source:HGNC Symbol;Acc:HGNC:18304] |
| Arl4c | ENSMUSG00000049866 | ARL4C | ENSG00000188042 | ADP ribosylation factor like GTPase 4C [Source:HGNC Symbol;Acc:HGNC:698] |
| Rpl27 | ENSMUSG00000063316 | RPL27 | ENSG00000131469 | ribosomal protein L27 [Source:HGNC Symbol;Acc:HGNC:10328] |
| Rrm1 | ENSMUSG00000030978 | RRM1 | ENSG00000167325 | ribonucleotide reductase catalytic subunit M1 [Source:HGNC Symbol;Acc:HGNC:10451] |
| Rpl30 | ENSMUSG00000058600 | RPL30 | ENSG00000156482 | ribosomal protein L30 [Source:HGNC Symbol;Acc:HGNC:10333] |
| Gbx2 | ENSMUSG00000034486 | GBX2 | ENSG00000168505 | gastrulation brain homeobox 2 [Source:HGNC Symbol;Acc:HGNC:4186] |
| Cux1 | ENSMUSG00000029705 | CUX1 | ENSG00000257923 | cut like homeobox 1 [Source:HGNC Symbol;Acc:HGNC:2557] |
| Dcaf12 | ENSMUSG00000028436 | DCAF12 | ENSG00000198876 | DDB1 and CUL4 associated factor 12 [Source:HGNC Symbol;Acc:HGNC:19911] |
| Klhl8 | ENSMUSG00000029312 | KLHL8 | ENSG00000145332 | kelch like family member 8 [Source:HGNC Symbol;Acc:HGNC:18644] |
| Zfy1 | ENSMUSG00000053211 | ZNF586 | ENSG00000083828 | zinc finger protein 586 [Source:HGNC Symbol;Acc:HGNC:25949] |
| Zfy1 | ENSMUSG00000053211 | ZNF480 | ENSG00000198464 | zinc finger protein 480 [Source:HGNC Symbol;Acc:HGNC:23305] |
| Zfy1 | ENSMUSG00000053211 | ZNF548 | ENSG00000188785 | zinc finger protein 548 [Source:HGNC Symbol;Acc:HGNC:26561] |
| Stag1 | ENSMUSG00000037286 | STAG1 | ENSG00000118007 | stromal antigen 1 [Source:HGNC Symbol;Acc:HGNC:11354] |
| Apc | ENSMUSG00000005871 | APC | ENSG00000134982 | APC regulator of WNT signaling pathway [Source:HGNC Symbol;Acc:HGNC:583] |
| Zfy1 | ENSMUSG00000053211 | ZNF549 | ENSG00000121406 | zinc finger protein 549 [Source:HGNC Symbol;Acc:HGNC:26632] |
| Zfy1 | ENSMUSG00000053211 | ZNF134 | ENSG00000213762 | zinc finger protein 134 [Source:HGNC Symbol;Acc:HGNC:12918] |
| Zfy1 | ENSMUSG00000053211 | ZNF304 | ENSG00000131845 | zinc finger protein 304 [Source:HGNC Symbol;Acc:HGNC:13505] |
| Zfy1 | ENSMUSG00000053211 | ZNF154 | ENSG00000179909 | zinc finger protein 154 [Source:HGNC Symbol;Acc:HGNC:12939] |
| Zfy1 | ENSMUSG00000053211 | ZNF793 | ENSG00000188227 | zinc finger protein 793 [Source:HGNC Symbol;Acc:HGNC:33115] |
| Zfy1 | ENSMUSG00000053211 | ZNF772 | ENSG00000197128 | zinc finger protein 772 [Source:HGNC Symbol;Acc:HGNC:33106] |
| Ptpn9 | ENSMUSG00000032290 | PTPN9 | ENSG00000169410 | protein tyrosine phosphatase non-receptor type 9 [Source:HGNC Symbol;Acc:HGNC:9661] |
| Hnrnpa1 | ENSMUSG00000046434 | HNRNPA1 | ENSG00000135486 | heterogeneous nuclear ribonucleoprotein A1 [Source:HGNC Symbol;Acc:HGNC:5031] |
| Myd88 | ENSMUSG00000032508 | MYD88 | ENSG00000172936 | MYD88 innate immune signal transduction adaptor [Source:HGNC Symbol;Acc:HGNC:7562] |
| Brd4 | ENSMUSG00000024002 | BRD4 | ENSG00000141867 | bromodomain containing 4 [Source:HGNC Symbol;Acc:HGNC:13575] |
| Ube2i | ENSMUSG00000015120 | UBE2I | ENSG00000103275 | ubiquitin conjugating enzyme E2 I [Source:HGNC Symbol;Acc:HGNC:12485] |
| Apc2 | ENSMUSG00000020135 | APC2 | ENSG00000115266 | APC regulator of WNT signaling pathway 2 [Source:HGNC Symbol;Acc:HGNC:24036] |
| Trh | ENSMUSG00000005892 | TRH | ENSG00000170893 | thyrotropin releasing hormone [Source:HGNC Symbol;Acc:HGNC:12298] |
| Pklr | ENSMUSG00000041237 | PKLR | ENSG00000143627 | pyruvate kinase L/R [Source:HGNC Symbol;Acc:HGNC:9020] |
| Krt18 | ENSMUSG00000023043 | KRT18 | ENSG00000111057 | keratin 18 [Source:HGNC Symbol;Acc:HGNC:6430] |
| Hnrnpr | ENSMUSG00000066037 | HNRNPR | ENSG00000125944 | heterogeneous nuclear ribonucleoprotein R [Source:HGNC Symbol;Acc:HGNC:5047] |
| Agrn | ENSMUSG00000041936 | AGRN | ENSG00000188157 | agrin [Source:HGNC Symbol;Acc:HGNC:329] |
| Cct3 | ENSMUSG00000001416 | CCT3 | ENSG00000163468 | chaperonin containing TCP1 subunit 3 [Source:HGNC Symbol;Acc:HGNC:1616] |
| Rpl26 | ENSMUSG00000060938 | RPL26 | ENSG00000161970 | ribosomal protein L26 [Source:HGNC Symbol;Acc:HGNC:10327] |
| Eef1b2 | ENSMUSG00000025967 | EEF1B2 | ENSG00000114942 | eukaryotic translation elongation factor 1 beta 2 [Source:HGNC Symbol;Acc:HGNC:3208] |
| Gfer | ENSMUSG00000040888 | GFER | ENSG00000127554 | growth factor, augmenter of liver regeneration [Source:HGNC Symbol;Acc:HGNC:4236] |
| Ephb2 | ENSMUSG00000028664 | EPHB2 | ENSG00000133216 | EPH receptor B2 [Source:HGNC Symbol;Acc:HGNC:3393] |
| Zfp454 | ENSMUSG00000048728 | ZNF454 | ENSG00000178187 | zinc finger protein 454 [Source:HGNC Symbol;Acc:HGNC:21200] |
| Rps20 | ENSMUSG00000028234 | RPS20 | ENSG00000008988 | ribosomal protein S20 [Source:HGNC Symbol;Acc:HGNC:10405] |
| Mllt11 | ENSMUSG00000053192 | MLLT11 | ENSG00000213190 | MLLT11 transcription factor 7 cofactor [Source:HGNC Symbol;Acc:HGNC:16997] |
| Pias4 | ENSMUSG00000004934 | PIAS4 | ENSG00000105229 | protein inhibitor of activated STAT 4 [Source:HGNC Symbol;Acc:HGNC:17002] |
| Rps13 | ENSMUSG00000090862 | RPS13 | ENSG00000110700 | ribosomal protein S13 [Source:HGNC Symbol;Acc:HGNC:10386] |
| Grwd1 | ENSMUSG00000053801 | GRWD1 | ENSG00000105447 | glutamate rich WD repeat containing 1 [Source:HGNC Symbol;Acc:HGNC:21270] |
| Ambra1 | ENSMUSG00000040506 | AMBRA1 | ENSG00000110497 | autophagy and beclin 1 regulator 1 [Source:HGNC Symbol;Acc:HGNC:25990] |
| Pik3ca | ENSMUSG00000027665 | PIK3CA | ENSG00000121879 | phosphatidylinositol-4,5-bisphosphate 3-kinase catalytic subunit alpha [Source:HGNC Symbol;Acc:HGNC:8975] |
| Rpl13 | ENSMUSG00000000740 | RPL13 | ENSG00000167526 | ribosomal protein L13 [Source:HGNC Symbol;Acc:HGNC:10303] |
| Gnb1 | ENSMUSG00000029064 | GNB1 | ENSG00000078369 | G protein subunit beta 1 [Source:HGNC Symbol;Acc:HGNC:4396] |

Supplementary Table 1. 558 unique mouse genes with absolute correlation index >= 0.5 were retained from GeneNetwork. These genes were converted to 381 unique human genes. The 381 unique human genes were filtered using BrainSpan data (overly expressed striatum genes in all prenatal vs adult samples) resulting in 72 genes. Of these 72 genes, 67 were found in GTEx and our study sample and were used to calculate striatum *SLC6A3 ePGS.*

**Supplementary Table 2**. Means of variables used in the cluster analysis for the ALSPAC cohort.

|  | Cluster - Low comorbidity risk |  | Cluster – High comorbidity risk |  | P-value |
| --- | --- | --- | --- | --- | --- |
|  | Mean / % | SE / N | Mean / % | SE / N |  |
| SDQ Total Difficulties | 5.02 | 0.14 | 6.77 | 0.26 | <0.001 |
| Depression Score | 0 | 0 | 1.08 | 0.06 | <0.001 |
| Anxiety Score | 0 | 0 | 0.88 | 0.06 | <0.001 |
| HOMA2-IR | 0.81 | 0.01 | 1.08 | 0.05 | <0.001 |
| Waist Circumference | 75.9 | 0.25 | 77.17 | 0.57 | 0.04 |
| Sex -Male | 49.5% | 434 | 42% | 131 | 0.03 |
| zBMI | .26 | 0.03 | 0.31 | 0.06 | 0.45 |
| Birth weight (kg) | 3.515 | 0.015 | 3.49 | 0.025 | 0.44 |
|  | N=876 |  | N=312 |  |  |

Supplementary Table 3. Main effect of PRS on psychiatric and cardio-metabolic comorbidity.

|  |  | ***Main effect*** | | |
| --- | --- | --- | --- | --- |
| **UK Biobank** | **PRS scores** | ***p*** | **β** | **OR** |
|  | Type 2 diabetes PRS_EPIC | < 0.001 | 0.073 | 1.076 |
|  | Major depression disorders PRS | < 0.001 | 0.065 | 1.068 |
| **ALSPAC** | **PRS scores** | ***p*** | **β** | **OR** |
|  | Type 2 diabetes PRS_EPIC | 0.253 | -0.076 | 0.927 |
|  | Major depression disorders PRS | 0.882 | 0.020 | 1.020 |

Supplementary Table 4. Interaction effect between PRS and birth weight on psychiatric and cardio-metabolic comorbidity.

|  |  | ***Interaction effect*** | | |
| --- | --- | --- | --- | --- |
| **UK Biobank** | **PRS scores** | ***p*** | **β** | **OR** |
|  | Type 2 diabetes PRS_EPIC | 0.490 | 0.014 | 1.014 |
|  | Major depression disorders PRS | 0.608 | -0.011 | 0.989 |
| **ALSPAC** | **PRS scores** | ***p*** | **β** | **OR** |
|  | Type 2 diabetes PRS_EPIC | 0.461 | 0.112 | 1.119 |
|  | Major depression disorders PRS | 0.798 | -0.078 | 0.925 |

| UK Biobank – Low vs randomly-selected Non-low Birth Weight group (n=1906) | | | | | |
| --- | --- | --- | --- | --- | --- |
| Characteristics | Low Birth Weight  (n =953) | | Non-low Birth Weight  (n = 953) | | P-value |
|  | Mean or  percentage | SD or N | Mean or  percentage | SD or N |  |
| Sex - Male | 34.7% | 331 | 44.7% | 426 | P<0.001 |
| Birth weight (grams) | 2129 | 376 | 3464 | 493 | P<0.001 |
| Completed full-time education at 14-years of age or younger | 0.7% | 4 | 0.9% | 5 | 0.97 |
| Age at recruitment (years) | 54.95 | 7.37 | 53.8 | 7.45 | P<0.001 |
| Townsend deprivation index at recruitment | -1.79 | 2.75 | -2.02 | 2.69 | 0.06 |
| BMI at recruitment | 26.83 | 4.45 | 26.52 | 4.24 | 0.11 |
| DAT1 STR ePGS | 0.02 | 0.95 | 0.07 | 0.96 | 0.29 |

**Supplementary Table 5.** Characteristics comparison between low vs randomly-selected non-low birth weight groups used in the parallel ICA analysis

**Supplementary Table 6.** Characteristics comparison between randomly-selected non-low birth weight group used in the parallel ICA analysis vs full sample of non-low birth weight individuals with MRI data available in the UK Biobank

| UK Biobank – randomly-selected non-low birth weight group vs rest of non-low birth weight group (n=10214) | | | | | |
| --- | --- | --- | --- | --- | --- |
| Characteristics | Randomly-selected Non-low Birth Weight  (n =953) | | Full sample of  Non-low Birth Weight  (n = 9261) | | P-value |
|  | Mean or  percentage | SD or N | Mean or  percentage | SD or N |  |
| Sex - Male | 44.7% | 426 | 42.3% | 3920 | 0.17 |
| Birth weight (grams) | 3464 | 493 | 3460 | 495 | 0.79 |
| Completed full-time education at 14-years of age or younger | 0.9% | 5 | 0.5% | 27 | 0.4 |
| Age at recruitment (years) | 53.8 | 7.45 | 53.75 | 7.38 | 0.85 |
| Townsend deprivation index at recruitment | -2.02 | 2.69 | -2.07 | 2.59 | 0.617 |
| BMI at recruitment | 26.52 | 4.24 | 26.51 | 4.34 | 0.956 |
| DAT1 STR ePGS | 0.07 | 0.96 | 0.05 | 0.96 | 0.607 |

Supplementary Table 7. Subset of significant SNPs related to gray matter density variations according to pICA analysis.

| Significant SNPs | |
| --- | --- |
| SNP | Z Score |
| rs34605159 | 12.99878 |
| rs7259029 | -11.3745 |
| rs139012629 | 11.11315 |
| rs115385848 | -9.25124 |
| rs116962884 | 8.970151 |
| rs11704200 | 8.799921 |
| rs184236146 | -7.28849 |
| rs6737722 | 7.180836 |
| rs112664980 | 5.614825 |
| rs56988084 | 5.071326 |
| rs2556378 | -5.00336 |
| rs76380377 | -4.92952 |
| rs67621412 | 4.895654 |
| rs72795692 | 4.635065 |
| rs6947066 | -4.32333 |
| rs1169557 | 4.131825 |
| rs3762272 | -3.79013 |
| rs372339543 | -3.7461 |
| rs58399787 | -3.63446 |
| rs2966318 | -3.50433 |
| rs2967848 | 3.478973 |
| rs140310740 | 3.390835 |
| rs70953651 | 3.35623 |
| rs149133487 | -3.20932 |
| rs71526493 | -3.17741 |
| rs3128100 | 3.123754 |
| rs2967871 | -3.06511 |
| rs117055030 | 2.941269 |
| rs1536096 | 2.915037 |
| rs11689362 | 2.813534 |
| rs3121575 | -2.7558 |
| rs57843539 | -2.61199 |
| rs11707190 | 2.509964 |
| rs2048008 | 2.500896 |
